# Improvements in clinical signs of Parkinson’s disease using photobiomodulation: A prospective proof-of-concept study

**DOI:** 10.1101/2021.05.26.21257833

**Authors:** Ann Liebert, Brian Bicknell, E-Liisa Laakso, Gillian Heller, Parastoo Jalilitabaei, Sharon Tilley, John Mitrofanis, Hosen Kiat

**Affiliations:** Department of Medicine and Health Sciences, University of Sydney, Camperdown, Australia; Governance and Research Department, Sydney Adventist Hospital, Wahroonga, Australia; Faculty of Health Sciences, Australian Catholic University North Sydney, Australia; Mater Research Institute, South Brisbane, Australia; Menzies Health Institute Queensland, Griffith University, Gold Coast, Australia; NHMRC Clinical Trials Centre, University of Sydney, Camperdown, Australia; Department of Mathematics and Statistics, Macquarie University, Macquarie Park, Australia; Department of Anatomy, University of Sydney, Camperdown, Australia; Lymphoedema And Laser Therapy, Stepney, Australia; Faculty of Medicine, Health and Human Sciences, Macquarie University, Australia; Faculty of Medicine, University of NSW, Kensington, Australia; Cardiac Health Institute, Sydney, Australia

**Keywords:** Parkinson’s disease, photobiomodulation, cognition, motor symptoms, mobility

## Abstract

**BACKGROUND:** Parkinson’s disease (PD) is a progressive neurodegenerative disease with no cure and few treatment options. Its incidence is increasing due to aging populations, longer disease duration and potentially as a COVID-19 sequela. Photobiomodulation (PBM) has been successfully used in animal models to reduce the signs of PD and to protect dopaminergic neurons.

**OBJECTIVE:** To assess the effectiveness of PBM to mitigate clinical signs of PD in a prospective proof-of-concept study, using a combination of transcranial and remote treatment, in order to inform on best practice for a larger randomized placebo-controlled trial (RCT).

**METHODS:** Twelve participants with idiopathic PD were recruited. Six were randomly chosen to begin 12 weeks of transcranial, intranasal, neck and abdominal PBM. The remaining 6 were waitlisted for 14 weeks before commencing treatment. After the 12-week treatment period, all participants were supplied with PBM devices to continue home treatment. Participants were assessed for mobility, fine motor skills, balance and cognition before treatment began, after 4 weeks of treatment, after 12 weeks of treatment and the end of the home treatment period. A Wilcoxon Signed Ranks test was used to assess treatment effectiveness at a significance level of 5%.

**RESULTS:** Measures of mobility, cognition, dynamic balance and fine motor skill were significantly improved (p<0.05) with PBM treatment for 12 weeks and up to one year. Many individual improvements were above the minimal clinically important difference, the threshold judged to be meaningful for participants. Individual improvements varied but many continued for up to one year with sustained home treatment. There was a demonstrable Hawthorne Effect that was below the treatment effect. No side effects of the treatment were observed.

**CONCLUSIONS:** PBM was shown to be a safe and potentially effective treatment for a range of clinical signs and symptoms of PD. Improvements were maintained for as long as treatment continued, for up to one year in a neurodegenerative disease where decline is typically expected. Home treatment of PD by the person themselves or with the help of a carer might be an effective therapy option. The results of this study indicate that a large RCT is warranted.

**TRIAL REGISTRATION:** Australian New Zealand Clinical Trials Registry, registration number: ACTRN12618000038291p, registered on 12/01/2018

## Introduction

Parkinson’s disease (PD) is the second most common neurodegenerative disorder after Alzheimer’s disease and the fastest growing neurodegenerative disease, due to an ageing population, a longer duration of the disease and possibly the increase in environmental contributors such as xenotoxins and environmental pollutants (1). It is also possible that the current COVID-19 pandemic may result in an increased incidence of PD in the future (2, 3). Deterioration in symptoms in sufferers of PD is the norm due to the progressive spread of α-synuclein mediated neuroinflammation, the loss of neurons in the substantia nigra and subsequent reduction in dopamine levels and decrease in mitochondrial function (4). To date there is no effective treatment that can cure or slow the progression of PD (5), although medications and deep brain stimulation can control some motor symptoms. The increasing recognition of the importance of the gut-brain axis in PD and the early presentation of gut symptoms (6), suggests the possibility of the gut as a target for PD therapies (7).

Photobiomodulation (PBM) therapy is the use of narrow-wavelength bands of non-thermal light (LED or laser) to modulate cellular responses. The main target of PBM is thought to be cytochrome-C-oxidase, which absorbs red and near-infrared light (8). This is thought to release reactive oxygen species (ROS) from the complex, promoting increased mitochondrial membrane potential, increased ATP production and regulate downstream cellular signalling pathways via ATP, cAMP, ROS, Ca^2+^ and nitric oxide (NO) to influences gene transcription (8, 9). PBM therapy has a decades-long safety record (10-12) with a safety profile equating to that of ultrasound tests. Unlike much pharmaceutical therapy, PBM therapy is free of serious deleterious side-effects and is non-invasive.

Because PBM acts at a cellular and mitochondrial level, the therapy has been shown to have a multitude of beneficial effects in the body and on various disorders, such as wound and diabetic ulcer healing, pain reduction, inflammatory disorders such as lung inflammation, osteoarthritis, tendinopathies and other musculoskeletal conditions (13, 14) In addition to the local effect of PBM on target cells, PBM also has a systemic effect (14-18) and a delayed effect due to activation of DNA transcription factors (8, 9). One of the primary downstream effects of PBM is on immune cells, producing an anti-inflammatory effect, which has profound consequences for many body processes (14). Recently there has been a great deal of interest in the use of transcranial PBM therapy to address symptoms of neurological and neuropsychiatric disorders (13).

Several studies have reported encouraging results for the application of PBM therapy in animal models of PD, and a recent review of animal evidence for treatment of PD with PBM concluded that human trials are justified (19). PBM has been shown to precondition and protect animals (including non-human primates) from a toxin (MPTP)-induced PD model, both in the signs of the induced PD and protection of the neurons in the substantia nigra (20-22). This preconditioning effect was also observed when PBM was delivered to areas remote from the brain (15, 23-25), including when the head was shielded from light (26). Several small trials and case studies are currently being undertaken with transcranial PBM (27-29). The application of remote PBM has not so far been investigated. In the current study, treatment consisted of combination of transcranial PBM and remote PBM treatment to the abdomen and to the neck was used. These locations were selected based on the importance of the gut-brain axis in PD, the richness of the enteric nervous system, the proximity of the vagus nerve in the neck and the success of these targets in animal models and clinical experience.

The aim of this proof-of-concept prospective clinical study was to assess the effectiveness of PBM to mitigate the clinical signs of PD in humans and to inform on treatment regimens and outcome measures for a future randomized placebo-controlled study (RCT). The primary outcome measure was improvement TUG as a measure of mobility. Secondary outcome measures were mobility, cognition, fine motor skill, micrographia and static balance. Quality of life outcome measures and patient reported symptomatic changes, including depression are the subject of a separate report.

## Methods

The study was conducted in Adelaide, Australia. The study received human research ethics approval by the Griffith University Human Research Ethics Committee (2018/16) and was registered with the Australian New Zealand Clinical trials Registry (ANZCTR - a primary registry in the WHO International Clinical Trial Registry Platform), registration number: ACTRN12618000038291p, registered on 12/01/2018. All participants gave written informed consent prior to taking part and all protocols were conducted in accordance with the ethics approval guidelines. The Consolidated Standards of Reporting Trials (CONSORT) guidelines were followed for this trial and a CONSORT flowchart (Figure 1A) summarizes participants treatments.

**Figure 1.**
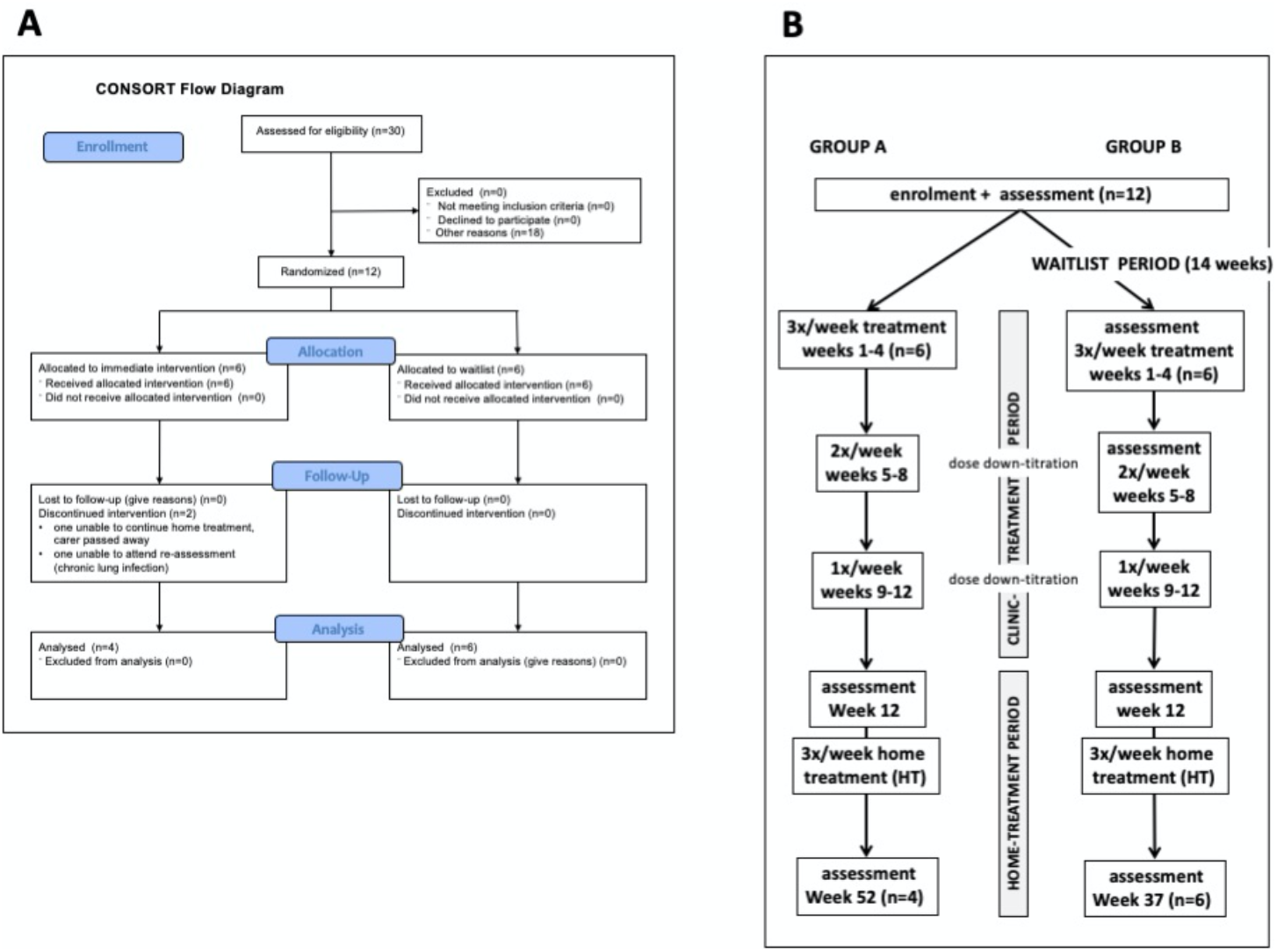
Study design: A. CONSORT flow chart of study design. B. Details of study design

### Study design

The study had a Waitlist Design (Figure 1B) with 6 participants (Group A) beginning treatment immediately in a clinic setting for 12 weeks (clinic-treatment period) beginning 14^th^ January 2019 and 6 participants (Group B) who acted as their own controls, beginning the same treatment protocol after a waiting period of 14 weeks (waitlist period), beginning 29^th^ July 2019. At the completion of the clinic-treatment period, all participants continued treatment with self-administered PBM devices at home for either 40 weeks (Group A) or 25 weeks (Group B).

### Participants

The participants in this study constituted a self-selected sample. Potential participants first rang the office of Parkinson’s South Australia (SA) in response to an advertisement in the Parkinson SA Newsletter and were given the list of inclusion and exclusion criteria. Inclusion criteria for the study were: age between 60 and 80 years, a diagnosis of idiopathic PD by a neurologist, Hoehn and Yahr stage I, II or II and a 6-month history of stable anti-PD medications (if taken). Exclusion criteria were: inability to self-care, a score of less than 24 on the Montreal Cognitive Assessment (MoCA) test, any psychotic episode or suicide ideation in the past 12 months, co-pathologies including uncontrolled cardiovascular disease, severe joint disease or orthopaedic injuries, peripheral neuropathy, vascular occlusive conditions, severe musculoskeletal conditions or vestibular conditions and any condition that would potentially interfere with PBM treatment such as structural brain disease, epilepsy or the use of potentially photosensitizing medication (e.g., imipramine, hypericum, phenothiazine, lithium, chloroquine, hydrochlorothiazide, tetracycline).

Individuals who fulfilled these criteria submitted a written application. The first 12 applicants were interviewed and examined by a neurologist who completed a MDS UPDRS assessment to ensure suitability for enrolment into the study. Participants were allocated into groups based on order of enrolment (Group A first 6; Group B subsequent 6).

### PBM treatment protocol

The PBM was administered transcranially with a VieLight Gamma device (4 LEDs, 240 joules), intranasally with a VieLight Gamma nasal device (1 LED, 15 joules), transdermally to the C1/C2 region of the neck and to the abdomen with an Irradia MID 2.5 laser device (4 laser diodes, 39.6 joules) or a MIDCARE laser device (2 diodes 39.6 joules). All participants received the same total energy dose from the PBM treatment throughout the study. Full PBM parameters are provided in Supplementary Table 1. The treatment protocol used LEDs and Class 1 lasers with no need for safety glasses.

The treatment during the clinic-treatment period was administered by a registered therapist, 3 times per week for weeks 1 to 4, reduced to twice per week for weeks 5 to 8, and further reduced to once per week for weeks 9 to 12 (dose down-titration). At the completion of the 12-week study period participants were supplied with treatment devices equivalent to those used in the 12-week treatment period (Supplementary Table 1). Participants and carers were given a 20-minute training session in the use of the equipment, which was essentially identical to the protocol that had been used for the past 12 weeks. The participants then continued self-administering the PBM treatment at home (home-treatment period) 3 times per week for an additional 40 weeks (Group A) or 25 weeks (Group B). Participants adherence to the treatment protocol was monitored by carers and reported at the final assessment.

### Participant Assessment

#### Safety

All participants were monitored for potential side effects of the PBM treatment during the 12-week clinic-treatment period. Participants were informed that a minority of people receiving PBM therapy can experience minor temporary side effects such as dizziness and/or mild nausea within 24 hours. Participants were questioned by the therapists on the second treatment and at weekly treatments thereafter to identify and assess any side effects. Participants and carers were instructed to address any concerns or perceived adverse reactions from the PBM treatment to the researchers or therapists during the clinic treatment and the home-treatment periods. Participants and carers reported on safety and side-effects at the final assessment.

#### Assessment

All participants were assessed before treatment began, after 12 weeks of treatment and after the home treatment period. Group B were additionally assessed before the waitlist period and after 4 weeks treatment (Figure 1B).

### Outcome Measures

The primary outcome measure (Table 1) was timed up-and-go (TUG), a measure of functional mobility (balance and mobility). Other outcome measures included additional tests of mobility (step test, TUG motor and TUG cognitive, walking speed and stride length), cognition, fine motor skill and static balance. All participants were assessed (Figure 1B) before treatment began, after the 12-week clinical-treatment period and after the home-treatment period (total PBM treatment of 52 weeks for Group A and 37 weeks for Group B). Group B was additionally assessed on enrolment into the study (14 weeks before treatment began) and after 4 weeks of treatment (before dose down-titration began). Assessments involving time were measured using stopwatch timers by 2 assessors and the mean time (seconds) was recorded to 2 decimal places.

**Table 1.**
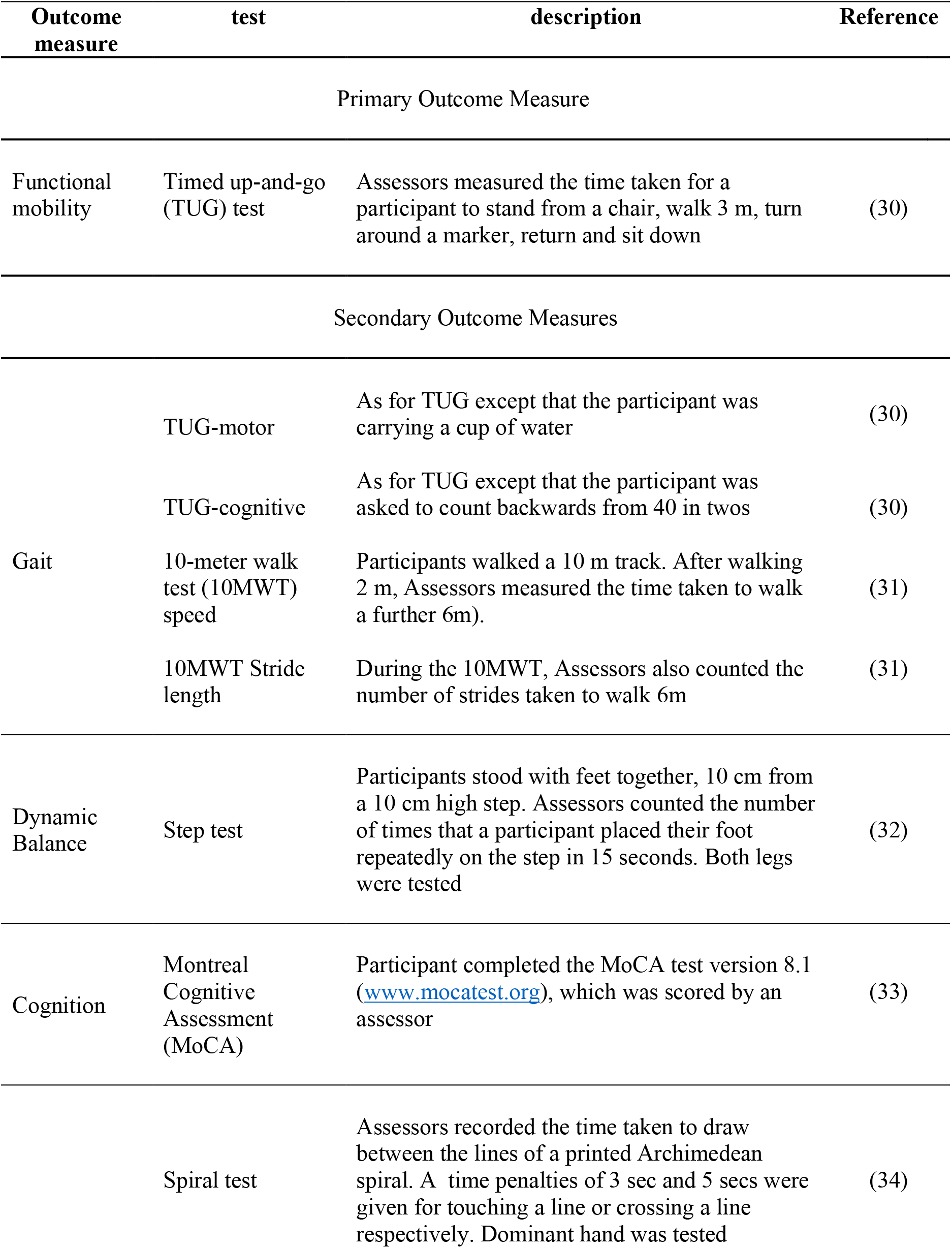
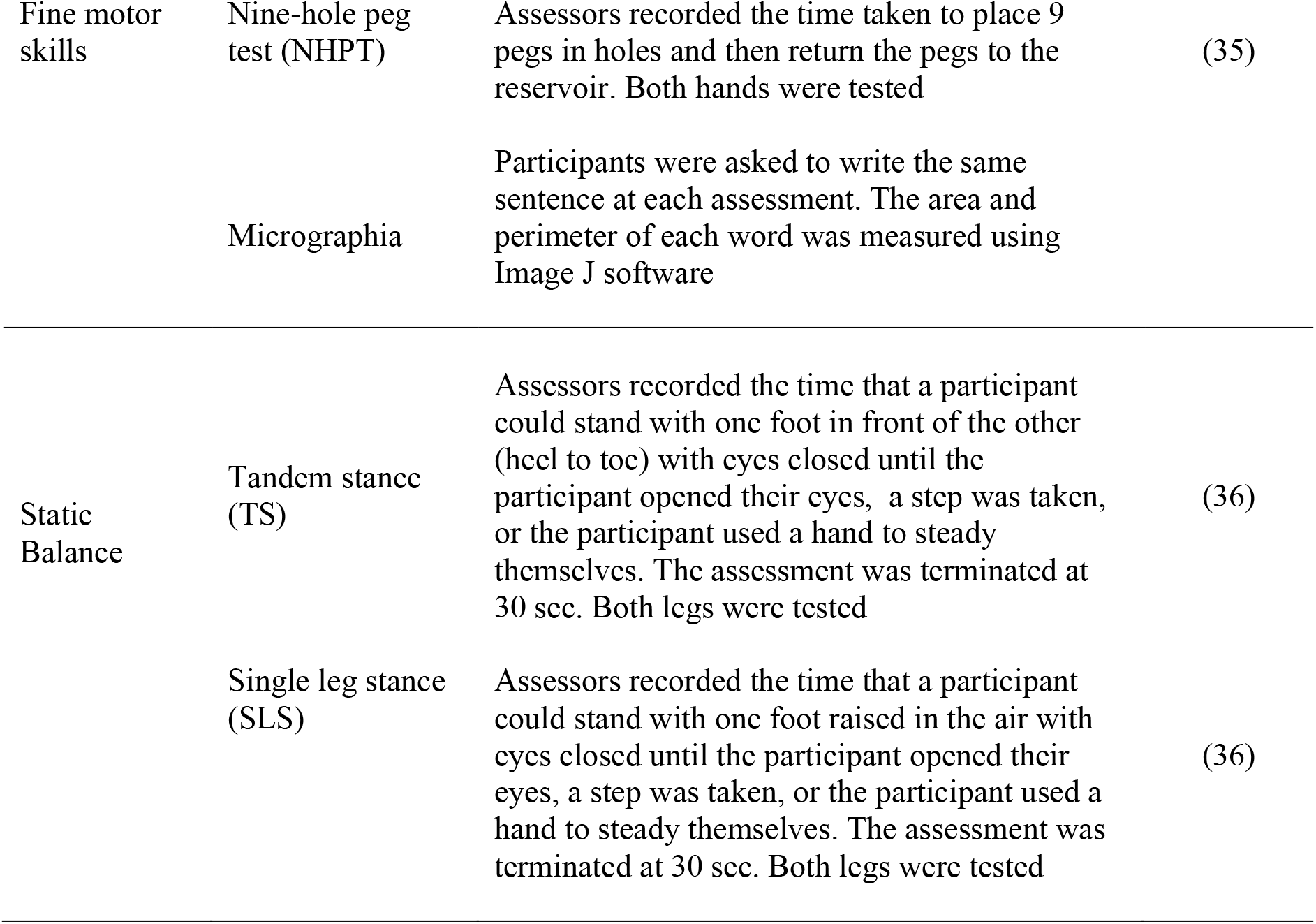
Outcome measures assessed before and after treatment with PBM

### Statistical analysis

Outcome measures were compared for paired data between assessment time points using the Wilcoxon Signed Ranks Test, since the sample size was judged to be too small for a complex statistical model such as analysis of variance (ANOVA). A significance level of 5% was used throughout with no adjustment for multiple comparisons such as a Bonferroni correction, since the reduction of Type I errors was considered to be of secondary importance in this exploratory proof-of-concept study. Two participants from Group A were not reassessed after the home-treatment period; one discontinued treatment and one had ongoing respiratory infection issues. Changes in outcome measures for individual participants were assessed using the Minimal Clinically Important Difference (MCID) with the change in an outcome measure compared to a baseline of the mean of all 12 participants, plus 7 participants from a second parallel proof-of-concept study (Supplementary Table 2). A participant was considered to have achieved a MCID improvement if the outcome measure improved by more than ½ standard deviation above the baseline (37). Larger changes were quantified as 1 and 2 standard deviations above the baseline measure.

## Results

### Participants

Participants were enrolled in the study in January 2019 and consisted of 7 females and 5 males, with an average age of 67.6 years and with a Hoehn and Yahr stage of between 1 and 3 (Table 2). Group A (immediate start to treatment) consisted of 4 males and 2 females with an average age of 71.3 years, of whom 5 (42%) were right-handed, and all were left side onset of PD (Table 2). Group B (waitlist group) consisted of 5 females and one male with an average age of 63.8 years, of whom 5 (42%) were right-handed and 3 (25%) were left side onset of PD.

**Table 2.** Summary of participant demographic characteristics

|  | Sex | Hoehn & Yahr stage | MDS UPDRS score | MDS UPDRS motor score | Dominant hand | Affected side |
| --- | --- | --- | --- | --- | --- | --- |
| A1 | M | 2 | 89 | 31 | R | L |
| A2 | F | 2 | 31 | 13 | R | L |
| A3 | F | 3 | 57 | 37 | R | L |
| A4 | M | 2 | 52 | 23 | R | L |
| A5 | M | 2 | 53 | 15 | R | L |
| A6 | M | 1 | 36 | 15 | L | L |
| B1 | F | 2 | 53 | 23 | R | L |
| B2 | F | 2 | 70 | 49 | R | L |
| B3 | F | 2 | 42 | 19 | R | R |
| B4 | M | 1 | 29 | 18 | R | L |
| B5 | F | 2 | 36 | 20 | L | R |
| B6 | F | 2 | 67 | 17 | R | R |

### Safety and Compliance

No adverse side effects or safety concerns were reported from the PBM treatment by the therapist, any participant or carer for the use of the therapy throughout the clinical-treatment and home-treatment periods. One participant (A2) suffered repeated respiratory infections during the home-treatment period and was unable to complete this part of the study and the assessment and a second participant received chemotherapy for breast cancer but continued with the study. Both of these occurrences were not considered to be side-effects of the PBM treatment protocol. 10 of 12 participants completed the home-treatment period of the study; A2 did not complete due to repeated respiratory infections and the carer of A3 passed away and she was unable to complete the treatment protocol. No participant reported a substantial change to their PD medications.

### Grouped outcomes

Full outcome measures data is available as Supplementary Table 3. Participants showed improvements in outcome measures (Table 3) after PBM therapy for up to 52 weeks. A statistically significant improvement (Wilcoxon Signed Ranks Test) was seen over the clinic-treatment period in the primary outcome measure of TUG and further improvement occurred over the home-treatment period. Secondary outcome measures that showed significant improvements over the clinic-treatment period included other tests of mobility (TUG motor, TUG cognitive, 10MWT walking speed and stride length), tests of balance (step test, TS test with affected leg behind), cognition (MoCA) and fine motor skill (spiral test). The step test and MoCA measures remained significantly improved above baseline after the period of self-administered home treatment (Table 4). No outcome measure showed a significant decline over the treatment period, although there was a non-significant increase in the median time to complete the NHPT.

**Table 3.** Medians (inter-quartile ranges) of outcome measures, on enrolment (before PBM treatment) and after PBM treatment for both groups.

|  | On enrolment<br>(n=12) | After 12 weeks of<br>clinic-treatment<br>(n=12) | After 25 or 40 weeks<br>of home-treatment<br>(n=10) |
| --- | --- | --- | --- |
| **Mobility tests** |  |  |  |
| 10MWT walk speed (m/s) | 1.12 (0.29) | 1.70 (0.35)** | 1.74 (0.41) |
| 10MWT stride length (m) | 0.52 (0.06) | 0.67 (0.11)** | 0.75 (0.06) |
| TUG (s) | 8.0 (1.6) | 7.1 (1.3)** | 6.58 (1.9)** |
| TUG motor (s) | 8.6 (3.2) | 7.6 (2.0)** | 10.4 (2.5)* |
| TUG cognitive (s) | 10.4 (2.5) | 6.9 (2.3)** | 9.5 (5.1)** |
| **Dynamic Balance test** |  |  |  |
| step test - affected leg (n) | 12.0 (5.0) | 16.5 (4.5)** | 17.5 (5.3)** |
| step test - unaffected leg (n) | 12.0 (2.0) | 15.5 (4.8)** | 19.5 (6.3)** |
| **Cognition test** |  |  |  |
| MoCA | 26 (3.0) | 28 (2.0)** | 29.9 (1.0)** |
| **Fine Motor Skill tests** |  |  |  |
| NHPT - affected hand (s) | 22.8 (4.0) | 27.5 (7.5) | 23.9 (7.0) |
| NHPT - unaffected hand (s) | 23.3 (6.0) | 24.0 (7.2) | 23.2 (6.2) |
| Spiral test - dominant hand (s) | 30.9 (11.0) | 27.3 (10.6)** | 23.4 (9.3)* n=11 |
| **Static Balance tests** |  |  |  |
| TS affected leg behind (s) | 2.0 (6.50) | 5.8 (17.00)* | 3.9 (26.6) n=9 |
| TS unaffected leg behind (s) | 1.5 (4.75) | 2.75 (19.08) | 3.9 (26.8) n=9 |
| SLS affected leg raised (s) | 2.0 (4.5) | 0.8 (3.3) | 4.5 (2.8) n=6 |
| SLS unaffected leg raised (s) | 1.5 (2.0) | 1.4 (3.4) | 24.5 (21.5) n=6 |

Significant improvement in outcome measure compared to before PBM treatment - \* $p < 0.05$ \*\* $p < 0.01$ 10MWT = 10 metre walk test; TUG = timed up-and-go; MoCA = Montreal Cognitive Assessment; NHPT = nine-hole peg test; TS = tandem stance (eyes closed); SLS = single leg stance (eyes closed)

**Table 4.** Medians (inter-quartile ranges) of outcome measures, before and after PBM treatment for group B (n=6).

|  | on enrolment | before PBM treatment | After 4 weeks PBM treatment | After 12 weeks PBM treatment |
| --- | --- | --- | --- | --- |
| **Mobility tests** |  |  |  |  |
| 10MWT walk speed (m/s) | 1.21 (0.24) | 1.82 (0.30)* | 1.94 (0.28)* | 1.93 (0.27) |
| 10MWT stride length (m) | 0.52 (0.09) | 0.67(0.11) | 0.71 (0.08) | 0.70 (0.08) |
| TUG (s) | 7.9 (1.6) | 7.4 (1.4)* | 6.6 (0.8)* | 6.9 (1.1) |
| TUG motor (s) | 8.6(1.2) | 8.2 (1.4) | 7.1 (0.7) | 6.9 (1.4) |
| TUG cognitive (s) | 9.6 (1.8) | 7.5 (1.1) | 7.0 (1.3)* | 6.8 (1.3) |
| **Dynamic Balance test** |  |  |  |  |
| step test - affected leg (n) | 10.0 (4.3) | 14.0 (2.0)* | 15.0 (2.3) | 15.5 (4.0) |
| step test - unaffected leg (n) | 11.5 (1.8) | 12.5 (3.3) | 15.0 (2.3) | 16.0 (5.0) |
| **Cognition test** |  |  |  |  |
| MoCA | 26.0 (2.3) | 27.5 (2.5) | 28.0 (1.5) | 29.0 (1.5) |
| **Fine Motor Skill tests** |  |  |  |  |
| NHPT - affected hand (s) | 24.6 (6.3) | 22.5 (7.0) | 24.0 (4.1) | 26.0 (9.7) |
| NHPT - unaffected hand (s) | 22.1 (2.1) | 25.4 (7.2) | 23.9 (4.7) | 24.0 (6.9) |
| Spiral test - dominant hand (s) | 35.2 (4.4) | 33.9 (10.9) | 27.3 (10.6) | 29.0 (6.2) |
| **Static Balance tests** |  |  |  |  |
| TS affected leg behind (s) | 2.0 (3.0) | 5.6 (6.2) | 19.5 (24.2)* | 5.8 (10.6) |
| TS unaffected leg behind (s) | 1.0 (3.0) | 4.2 (7.0) | 3.2 (20.3) | 2.3 (13.6) |
| SLS affected leg raised (s) | 1.5 (1.8) | 1.6 (1.0) | 1.2 (1.2) | 0.8 (4.5) |
| SLS unaffected leg raised (s) | 1.0 (0.8) | 2.1 (1.8) | 0.1 (1.2) | 0.9 (4.8) |

Significant improvement in outcome measure - \* p<0.05
10MWT = 10 metre walk test; TUG = timed up-and-go; MoCA = Montreal Cognitive Assessment;
NHPT = nine-hole peg test; TS = tandem stance (eyes closed); SLS = single leg stance (eyes closed)

For Group B, there was an improvement in many outcome measures during the waitlist-period (Table 4), which was significant for walk speed and TUG. Apart from NHPT, the improved outcomes further improved after PBM treatment commenced. For walk speed, TUG, TUG cognitive and tandem balance (with affected foot back) these were significant improvements. Dose down-titration after week 4 resulted in reduction in many of the improvements made by participants in Group B (Table 4), most notably TS with the affected leg back. Despite the dose down-titration over 8 weeks, some outcome measure continued to improve (most notably cognition and the step test. The improvements in mobility, cognition and spiral test attained in week 4 were sustained until assessment at week 12.

There was no significant change in participants’ handwriting and the area and perimeter of words between the three assessment times in either group (Figure 2). The results for one participant were incomplete and could not be included in the analysis.

**Figure 2.**
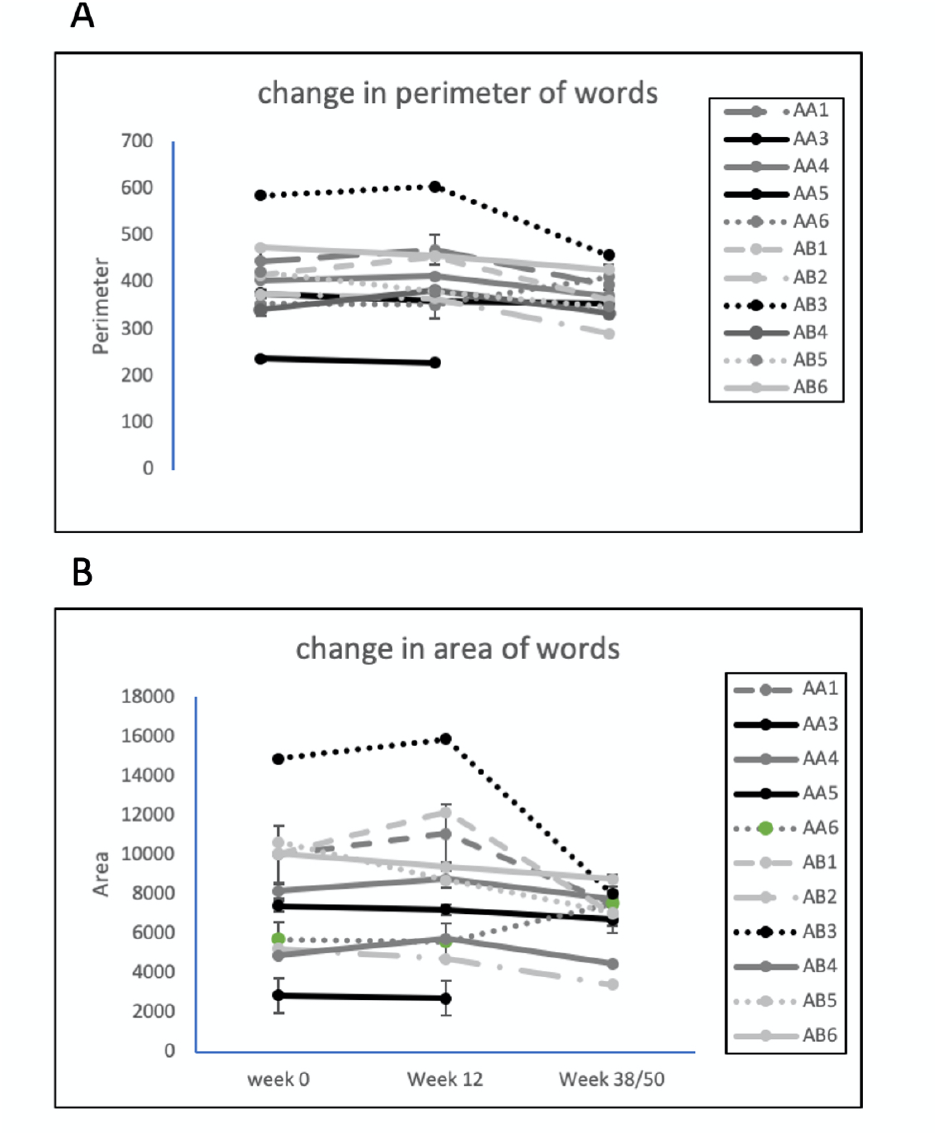
Changes in micrographia of participants over the photobiomodulation treatment period. A – perimeter of words; B – area of words. Group A: n=5, P = 0.95, F= 0.05; Group B: n=6, P = 0.24, F= 1.6.

### Individual data

Changes to individual participant outcome measures are displayed as heatmaps (Figure 3) to represent changes from baseline. Multiple outcome measures improved for all participants over the 12-week clinic-treatment period (Figure 3a), many of which were equivalent to or better than a MCID. For all participants, improvements persisted during the home-treatment period (Figure 3b) for walk speed, TUG, step test, the MoCA and the spiral test. Some outcome measures, such as MoCA, continued to improve during the home-treatment period. The least improved outcome measures were the NHPT and static balance (TS and SLS). Group B participants showed improvement in 64 of the 90 outcome measures assessed during over the waitlist period (Figure 3c). After treatment began, 51 of these 64 outcome measures showed further improvement (at the 4-week assessment) while 9 declined. The most notable improvement after treatment commenced was in TS (Table 4), which was also the outcome measure that showed the greatest decline after dose down-titration (12-week assessment).

**Figure 3.**
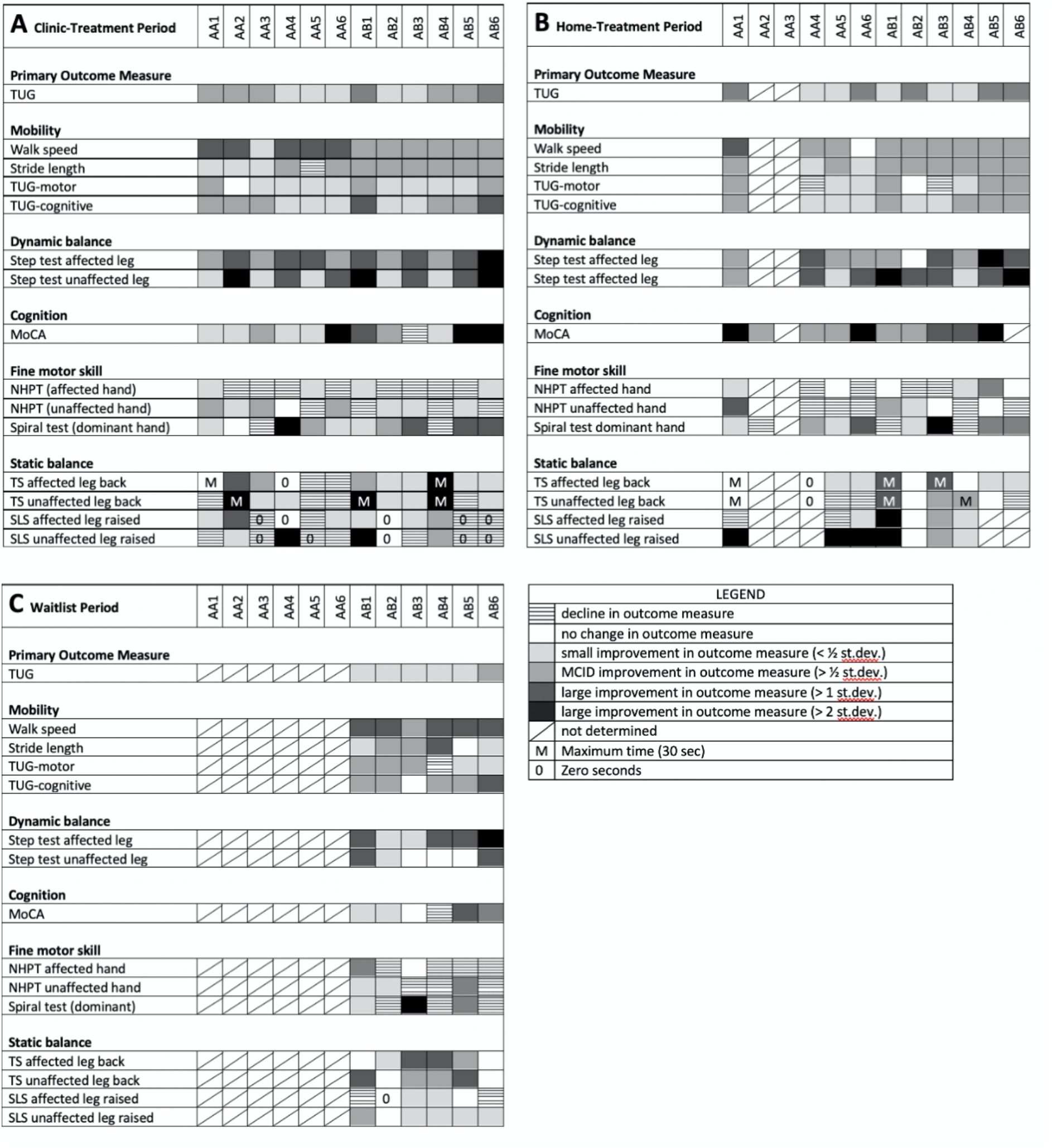
Heatmap depicting changes in outcome measures after PBM treatment, compared to enrolment: (A) after 12 weeks of PBM treatment in a clinic setting; (B) after 37 or 52 weeks of PBM treatment (clinic-treatment + self-administered home-treatment) ; (C) after 14 weeks of waitlist with no treatment. The columns are individual participants. The rows are assessed outcome measures. Shades of grey represent improvements in outcome measure; no colour represents no change in outcome measure; hatched represents a decline in an outcome measure; a diagonal bar represents no data for the outcome measure.

Outcome measure assessment was influenced by individual circumstances of the participants. In addition to the cancer therapy of one participant and the respiratory infections of a second, the partner of a third participant passed away during the trial and the participant was unable to complete the self-administered home treatment protocol and the carer of a fourth participant withdrew from their domestic relationship during the clinic-treatment period.

## Discussion

We have shown that PBM treatment is capable of improving a number of clinical signs of Parkinson’s disease, including the primary outcome measure of TUG, which assesses functional mobility, and also other mobility related signs, some fine motor skills and cognition. These improvements persisted for up to one year with continued PBM treatment. Importantly, there was no significant decline in any outcome measure over one year, although there were small (non-significant) declines in the NHPT and in micrography. To the best of our knowledge, the study described herein represents the first clinical trial in PD patients using PBM treatment to a combination of anatomical targets, although several small trials and case studies using transcranial PBM for PD are currently underway with (27-29). Although the study reported here was not sufficiently powered to detect irrefutable changes in clinical signs of PD, the results build on results from animal studies and demonstrate the potential clinical relevance of the PBM treatment in mitigating clinical signs of PD. Importantly the treatment presented no safety concerns and the participants reported no adverse side-effects, confirming the safety of PBM as has been seen in numerous other studies of PBM treatment

PD currently has no cure and there are few options to arrest or slow the signs and symptoms of the disease and so treatment is based on symptomatic relief. The gold standard for treatment is dopamine replacement with levodopa, combined with carbidopa to prevent premature conversion to dopamine. These can improve motor symptoms but can also cause adverse side effects, such as dyskinesia and nausea, and become less effective with time. Other clinically useful medications for motor symptoms include dopamine agonists, ergot, MAO-B inhibitors, anticholinergics and Adenosine agonists, which can be used alone or as adjunct therapy (5). Because the signs and symptoms of PD are diverse, the pharmacological treatment of the mixture of symptoms is challenging and often necessitates a cocktail of pharmacologic interventions (38), depending on individual patient needs (39, 40).

Recent evidence-based review of treatment options for motor and non-motor signs and symptoms of PD, commissioned for the International Parkinson’s and Movement Disorder Society (5, 41) concluded that there are few non-pharmaceutical options to control motor symptoms. Of these, exercise and physiotherapy are common interventions that are clinically useful, while supplements (e.g., coenzyme Q_10_, creatine, Vitamin D), lack clinical evidence despite being popular with PD sufferers (5). Deep brain stimulation is an established surgical technique that controls some motor symptoms (stiffness, tremor) and can improve quality of life, but like all surgery carries some risk (42). For non-motor symptoms, it was concluded that “There were no clinically useful interventions identified to treat non-dementia-level cognitive impairment.” although there were some pharmacological options for dementia (41). A number of new interventions for PD are currently undergoing investigation, including high intensity focussed ultrasound (43), immunotherapy (44) and stem cell therapy (45). The current study provides early clinical evidence that PBM has the potential to be an effective treatment complimenting traditional pharmaco- and physical therapy in the management of the clinical signs of PD.

The primary outcome measure of the study was functional mobility as measured by TUG. Not only was this outcome measure significantly improved after the 12-week clinic-treatment period and the home-treatment period, but all participants showed this improvement. Motor symptoms of PD have a major impact on the quality of life of PD sufferers (46) and are complex, being a combination of mobility, balance and cognition. All measures of mobility were improved in all participants, with significant improvements in walking speed, stride length, step test and TUG tests throughout the clinic-treatment and home-treatment periods. Both the 10MWT and the three TUG tests are validated for PD (31, 47), show good reliability and relationship to mobility, falls risk and the progression of disease (48). Increased falls risk is also related to loss of balance, severity of PD and previous falls history (49). Although the step test was originally developed for stroke patients and includes a component of physical capacity, it is simple to perform and has found some utility for PD patients (36, 50). Participants in the current study showed significant improvement in the step test, with improvements being maintained through the home-treatment period. On the other hand, the TS and SLS tests of static balance, although somewhat improved with PBM, were also the most sensitive to dose down-titration. Mobility dysfunction is also related to cognition and the ability to integrate sensory information and motor planning (51) as demonstrated by dual-task TUG (TUG-motor and TUG-cognitive) in PD (52). Improvements in these outcome measures have the potential to positively impact the mobility of individuals with PD and so reduce the risk of falls.

Another notable outcome of the study was the improvement in cognition as assessed by the MoCA, especially considering that up to 80% of PD patients develop dementia within 15–20 years of onset (53). The MoCA is considered a suitable cognitive assessment screening tool in PD (54) and has excellent test-retest reliability with no significant learning effects, even when used within one month (www.mocatest.org). Supporting the MoCA outcomes were anecdotal comments by study participants and carers who remarked on improvements in mood, engagement and socialisation (data not shown). In a recent review of treatment options for non-motor signs and symptoms of PD (41), it was concluded that “there were no clinically useful interventions identified to treat non-dementia-level cognitive impairment”. There have been a number of previous reports of improved cognition using transcranial PBM (55-57), often in conjunction with intranasal PBM (58), including the VieLight device used in the current study (59).

MDS UPDRS was not used for assessment of outcomes in this study, due to the unavailability of the consulting neurologist at various stages of the study. While the MDS UPDRS is recognised as the “Gold Standard” for PD diagnosis, it may lack sensitivity to detect changes in the signs and symptoms in early PD and its progression (60-62), especially for functional performance (63) and cognition (64). It remains to be seen if the UPDRS would be suitable to detect improvements in the clinical signs of PD that were seen with PBM treatment in the current study.

Many of the participants improved in multiple outcome measures as measured by improvement above an MCID value. MCID is interpreted as an improvement that is relevant to individual participants (65) and is proposed as the “smallest difference in score in the domain of interest which patients perceive as beneficial and which would mandate, in the absence of troublesome side effects and excessive cost, a change in the patient’s management” (66). The use of a half standard deviation as a simple measure of MCID as proposed by Norman et al (37) is not universally accepted (67) and its use in our study with its small number of participants has resulted in a standard deviation that is higher than would be expected with a larger cohort, leading to imprecision in detecting an MCID change, with an MCID change more difficult to achieve, and an underestimate of the numbers of participants that show a substantial improvement. This is most apparent in the measures of static balance (TS and SLS), where the high variance resulted in few participants achieving an MCID, despite some substantial improvements. While less than ideal, the categorizing of improvements in clinical signs as MCID provides a consistent although high benchmark against which to measure improvements. At a time when the consumer perspective is considered imperative in clinical research, especially in a disease with such diverse symptoms as PD, future studies using a measure of MCID would ideally be related to participant’s perceptions of what they themselves considered an ‘important difference’ to their own clinical signs and symptoms.

It was apparent that there was considerable variability among participants in response to the PBM treatment, which is important to consider at this early stage of clinical study. While all participants showed improvement in multiple clinical signs, the number and specific sign, as well as the extent of the improvement varied among participants. Many participants showed an improvement after PBM treatment for a range of outcome measures (e.g., A1, A4, B1, B2, B5, B6), while some participants showed an improvement in fewer outcome measures (e.g., A5).This is not unexpected due to the variability of signs and symptoms among PD patients and the heterogeneity of this small participant group. Variability in individual response to the PBM treatment may also be due to individual responses to light in general and to PBM in particular (68). A number of personal and unavoidable circumstances may also have adversely influenced performance of assessments after the clinic-treatment and home-treatment periods. Future studies will need to take account of the variability in the symptomology of PD participants and enrol sufficient numbers into the study to ensure statistical power to demonstrate improvements in clinical signs and symptoms.

The most noteworthy individual result was the maintenance of improvement in some outcome measures for up to one year with the continued self-administered home treatment. Indeed, some outcome measures such as the MoCA improved further during the home-treatment period and few of the outcome measures declined over this period. As a neurodegenerative disease in which motor and non-motor function would be expected to gradually decline, the improvement with PBM treatment in some of the clinical signs of PD and the preservation of this improvement over time is clinically relevant and worthy of further validation in longer term trials. Longer term treatment in the home setting also appears to be a practical and cost-effective strategy, with the treatment performed by the participant with or without the help of a carer. A second noteworthy result was the diminution of improvement with down-titration of the PBM dose, which resulted in a reversal of some of the most marked improvements at week 4 (such as TS). Dose down-titration is a common strategy in PBM therapy, developed by Chow (personal communication) among others, based on protocols for the relief of pain (69). This strategy can inform on the most effective dose of PBM and whether the therapy is able to be reduced or withdrawn, as is the case with pain management and wound healing with PBM. The reversal of the improvements with dose down-titration suggests that the PBM treatment needs to be maintained at a suitable level and that the dosing regimen is central to maximising treatment success. This observation informed on the dose regimen of the home-treatment (3 times per week). Despite the diminution of improvement after week 4, outcome measures remained significantly improved over baseline for the majority of outcome measures during the 12-week clinic-treatment period.

Micrographia is a common and often early sign of PD and can overlap with other signs and symptoms of PD (70). There was no significant change in participants’ handwriting during the PBM treatment period, which might indicate a stabilisation rather than the expected decline in participant’s micrographia. The stabilization of handwriting was also noted in a case series using transcranial PBM (28), with 6 of 6 PD participants showing no decline over 24 months. Dopaminergic therapy and deep brain stimulation have not been shown to slow the decline in writing size (71).

The placebo effect can be pronounced in clinical trials and is well known in PD therapy (72). The current study did not have a placebo arm to quantify the placebo effect, but the related Hawthorne Effect could be assessed. The Hawthorne Effect can occur in response to participation in research or being observed during a study (73, 74) and has been recognised as a confounder to results in clinical trials of PD (75), such as the evaluation of gait being affected (not significantly) by whether the participant was observed overtly or covertly (76). The Hawthorne Effect appears to be transient, being short-lived during the treatment period (75) and much diminished by 3 months (77). In the current study, the waitlisted participants (Group B) showed an improvement in outcome measures before treatment began, with some of these improvements being sufficient to qualify as an MCID, thus demonstrating a measurable Hawthorne Effect. The other possibility is that the participants have improved due to a practice effect with the repeated assessments. While possible, it is unlikely to completely explain the improvement during the 14-week period between enrolment and treatment. A practice effect cannot be entirely excluded for subsequent improvement at the 4-week assessment after treatment began and is a potential confounder for this assessment. PD patients have been shown to have a diminished ability for motor learning and require increased practice sessions for balance related tasks, compared to young healthy controls (78).

While a placebo, Hawthorne or a practice effect as the sole explanation for all improvements seen in this study cannot be entirely ruled out, it appears quite unlikely. Most outcome measures showed continued and accelerated improvement once treatment began (Table 4) and the improvements in outcome measures were maintained throughout the home-treatment period (Table 3, Figure 3b). The Hawthorne and placebo effects would be expected to be transitory and would, at the minimum, diminish during the home-treatment period when there was no continued interaction with study therapists and researchers. A more thorough randomized placebo-controlled trial is warranted to more fully explore the placebo effect in treatment of PD by PBM.

The mode of action of PBM treatment in PD merits further research. Transcranial PBM has recently been assessed for its effectiveness for a number of brain-related conditions and injuries including stroke (79), traumatic brain injury (55), post-traumatic stress disorder (80), depression (81) and Alzheimer’s disease (59). Transcranial devices have been shown to modulate neural oscillations (82, 83). A transcranial device has also been used as a treatment for PD in a series of case studies (28, 84) with encouraging results, especially for non-motor symptoms. Evidence from experimental and animal models suggests that transcranial PBM could act via the cytochrome-C-oxidase target of near infrared light, to increase ATP and influence downstream cellular signalling to reduce oxidative stress and neuroinflammation and to upregulate synaptogenesis and neurogenesis (85).

Treatment of areas remote from the injury/disease has been shown to be an effective therapy strategy. For example, targeting the tibia with PBM can help in the repair of cardiac tissue (17). The mechanism of this systemic effect of PBM is likely to be pluralistic, through the stimulation of stem cells (24, 86), an immune modulation response (87), circulating cell-free functional mitochondria (88), or by circulating chemical messengers (23). The use of the 904nm laser PBM protocol was based on experimental models of remote PD treatment by various wavelengths of LED and laser in a mice model (unpublished data).

The study reported here is the first to use a combination of transcranial and remote abdominal PBM treatment for PD. The abdomen is an appropriate target for remote application of PBM for PD, given the strong gut-brain axis link for the disease and previous results of remote application in animal model studies (23, 24). The combination of PBM treatments used in the current study improved mobility and other clinical signs and symptoms of PD, including cognition, possibly by compensating for the loss of neuronal connections caused by the progressive lack of dopamine. Further study is required to ascertain the optimal sites of treatment, the optimal dose regimen and the precise mechanism of action.

## Conclusion

To the best of our knowledge the study described here represents the first clinical trial in people with PD using PBM treatment to a combination of anatomical targets. PBM was shown to be a safe, side-effect free, low cost and a potentially clinically effective treatment, with significant improvements in primary and secondary outcome measures. Improvements to participant outcome measures were maintained for up to one year with continued treatment, which is a persuasive indication of the effect of PBM treatment. The results suggest that PBM treatment may slow some of the expected decline in the clinical signs and symptoms of PD and may complement the current treatment options. The full potential of PBM as an intervention for the signs and symptoms of PD needs to be further scrutinized and validated in a larger, prospective, randomized placebo-controlled trial with sufficient power and a longer follow-up period.

## Data Availability

All data generated and analysed during this study for this report are included in this published article and its supplementary information files. Additional study data can be requested from the corresponding author on request.

## Abbreviations

10MWT: 10 metre walk test
ANOVA: analysis of variance
ANZCTR: Australian New Zealand Clinical Trials registry
ATP: adenosine triphosphate
cAMP: cyclic adenosine monophosphate
CONSORT: Consolidated Standards of Reporting Trials
COVID: coronavirus disease of 2019
LED: light emitting diode
MCID: minimal clinically important difference
MDS UPDRS: Movement Disorder Society Unified Parkinson’s Disease Rating Scale
MoCA: Montreal cognitive assessment
MPTP - 1: methyl-4-phenyl-1,2,3,6-tetrahydropyridine
NHPT: nine-hole peg test
NO: nitric oxide
PD: Parkinson’s disease
PBM: photobiomodulation
RCT: randomized placebo-controlled trial
ROS: reactive oxygen species
SA: South Australia
SLS: single leg stance
TS: tandem stance
TUG: timed up and go
WHO: World Health Organisation

