## Supplementary Table 1 for "Improvements in clinical signs of Parkinson’s disease using photobiomodulation: A prospective proof-of-concept study"

**Supplementary Table 1.** Parameters of the photobiomodulation devices and treatment used in the study.

| PARAMETER | SYMBYX PDCARE LASER | | VIELIGHT NEURO GAMMA | | |
|---|---|---|---|---|---|
| Manufacturer | Spectra Analytic Irradia AB | | Vielight Inc. | | |
| Diodes | 904nm laser diodes (GaAs) | | 5 x LED diodes | | |
| Wavelength | 904nm | | 810 nm | | |
| Laser class | 1 | | - | | |
| | Clinic laser | Home use laser | posterior | anterior | nasal |
| Number of diodes | 4 | 3 | 3 | 1 | 1 |
| Output power | 30 mW | 30 mW | 100 mW | 75 mW | 25mW |
| Peak power | 25,000 mW | 25,000 mW | | | |
| Pulse frequency | 50 Hz | 50 Hz | 40 Hz | 40 Hz | 40 Hz |
| Beam spot size | 0.635 $cm^2$ | 0.635 $cm^2$ | ~1 cm | ~1 cm | ~1 cm |
| Power density per diode | 47 $mW/cm^2$ | 47 $mW/cm^2$ | 100 $mW/cm^2$ | 75 $mW/cm^2$ | 25 $mW/cm^2$ |
| Total output power | 120 mW | 60mW | 400 mW | | |
| Irradiation time per point | 30 s | 60 s | 2100 s | | |
| Total irradiation time | 330 s | 660 s | 2100 s | | |
| Total energy per point | 3.6 J | 3.6 J | 60 J | 45 J | 15 J |
| Number of sites | 11 (9 abdomen, 2 neck) | 11 (9 abdomen, 2 neck) | 3 | 1 | 1 |
| Total energy dose per treatment | 39.6 J | 39.6 J | 180 J | 45 J | 15 J |
| Treatment frequency weeks 1 to 4 | 3 x per week for 4 weeks | | 3 x per week for 4 weeks | | |
| weeks 5 to 8 | 2 x per week for 4 weeks | | 2 x per week for 4 weeks | | |
| weeks 9 to 12 | 1 x per week for 4 weeks | | 1 x per week for 4 weeks | | |
| weeks 12 to 37 or 52 | | 3 x per week for 25 or 40 weeks | 3 x per week for 25 or 40 weeks | | |
