## Supplementary Table 2 for "Improvements in clinical signs of Parkinson’s disease using photobiomodulation: A prospective proof-of-concept study"

**Supplementary Table 2**. Mean (st dev) of measures of fine motor control, gait, balance and cognition before PBM treatment in a second study of PD treatment with PBM

| | Before treatment |
|---|---|
| **Gait tests** | |
| 10MWT walk speed (m/sec) | 1.69 (0.51) n=7 |
| 10MWT stride length (m) | 0.97 (0.24) n=7 |
| TUG (s) | 7.2 (2.42) n=7 |
| TUG motor (s) | 8.1 (1.85) n=7 |
| TUG cognitive (s) | 8.4 (2.33) n=7 |
| **Dynamic Balance test** | |
| Step test - affected leg (n) | 15.3 (4.03) n=7 |
| Step test - unaffected leg (n) | 16.6 (5.02) n=7 |
| **Cognition test** | |
| MoCA | 25.7 (3.08) n=6 |
| **Fine Motor Skill tests** | |
| NHPT - affected hand (sec) | 29.9 (7.60) n=6 |
| NHPT - unaffected hand (sec) | 26.4 (4.79) n=6 |
| Spiral test - dominant hand (sec) | 25.6 (9.78) n=6 |
| **Static Balance** | |
| TS affected leg behind (sec) | 14.9 (17.70) n=7 |
| TS unaffected leg behind (sec) | 13.0 (10.94) n=7 |
| SLS affected leg raised (sec | 7.4 (10.25) n=7 |
| SLS unaffected leg raised (sec) | 8.9 (10.59) n=7 |

10MWT = 10 metre walk test; TUG = Timed-up-and-go; MoCA: Montreal Cognitive Assessment; NHPT = nine-hole peg test; TS = tandem stance (eyes closed); SLS: single leg stance (eyes closed)
