## Supplementary Table 3 for "Improvements in clinical signs of Parkinson’s disease using photobiomodulation: A prospective proof-of-concept study"

**Supplementary Table 3. Individual data for participants before and after treatment with the photobiomodulation protocols.**

| | GROUP AA PARTICIPANTS | | | | | | GROUP AB PARTICIPANTS | | | | | |
|---|---|---|---|---|---|---|---|---|---|---|---|---|
| | AA1 | AA2 | AA3 | AA4 | AA5 | AA6 | AB1 | AB2 | AB3 | AB4 | AB5 | AB6 |
| **NHPT (seconds)** | | | | | | | | | | | | |
| **affected hand** | | | | | | | | | | | | |
| 12 weeks prior to treatment | | | | | | | 29.0 | 45.8 | 19.6 | 25.1 | 21.0 | 24.0 |
| immediately before treatment | 22.5 | 23.0 | 21.2 | 26.1 | 21.5 | 21.6 | 22.2 (23%) | 56.5 -(23%) | 19.5 (1%) | 31.3 -(24%) | 22.3 -(6%) | 22.8 (5%) |
| after 4 weeks treatment (CT) | | | | | | | 25.8 (11%) | 48.2 -(5%) | 21.5 -(10%) | 25.7 -(2%) | 22.3 -(6%) | 19.2 (20%) |
| after 12 weeks treatment (CT) | 21.1 (6%) | 28.6 -(24%) | 28.0 -(32%) | 37.1 -(42%) | 17.9 (17%) | 27.1 -(25%) | 27.9 (4%) | 47.6 -(4%) | 20.5 -(4%) | 34.8 -(38%) | 24.1 -(15%) | 23.2 (3%) |
| after 26/38 week home treatm | 21.8 (3%) | | | 27.3 -(5%) | 21.6 (0%) | 29.6 -(37%) | 29.4 -(1%) | 69.9 -(53%) | 22.0 -(12%) | 23.4 (7%) | 16.4 (22%) | 24.4 -(2%) |
| **dominant hand** | | | | | | | | | | | | |
| 12 weeks prior to treatment | | | | | | | 20.1 | 30.2 | 19.6 | 24.1 | 21.9 | 24.0 |
| immediately before treatment | 29.3 | 18.1 | 33.7 | 26.4 | 16.0 | 25.7 | 18.0 (11%) | 29.1 (4%) | 19.5 (1%) | 26.6 -(10%) | 17.9 (18%) | 22.8 (5%) |
| after 4 weeks treatment (CT) | | | | | | | 16.5 (18%) | 24.9 (18%) | 21.5 -(10%) | 26.8 -(11%) | 18.7 (15%) | 19.2 (20%) |
| after 12 weeks treatment (CT) | 25.8 (12%) | 17.9 (1%) | 27.7 (18%) | 26.8 -(2%) | 17.7 -(10%) | 20.0 (22%) | 25.5 -(27%) | 28.3 (6%) | 20.5 -(4%) | 28.8 -(20%) | 20.1 (8%) | 23.2 (3%) |
| after 26/38 week home treatm | 21.8 (26%) | | | 27.9 -(6%) | 17.0 -(6%) | 27.1 -(5%) | 17.1 (15%) | 29.6 (2%) | 20.1 -(2%) | 28.8 -(20%) | 21.4 (2%) | 22.5 (6%) |
| **SPIRAL TEST (seconds)** | | | | | | | | | | | | |
| **dominant hand** | | | | | | | | | | | | |
| 12 weeks prior to treatment | | | | | | | 32.4 | 37.5 | 48.6 | 33.9 | 29.3 | 36.5 |
| immediately before treatment | 20.4 | 23.8 | 29.3 | 52.5 | 22.6 | 26.2 | 29.6 (9%) | 38.2 -(2%) | 28.7 (41%) | 42.0 -(24%) | 20.7 (29%) | 40.3 -(10%) |
| after 4 weeks treatment (CT) | | | | | | | 19.4 (40%) | 30.8 (18%) | 18.7 (61%) | 41.7 -(23%) | 23.7 (19%) | 31.2 (15%) |
| after 12 weeks treatment (CT) | 16.8 (18%) | 23.7 (0%) | 31.9 -(9%) | 23.2 (56%) | 16.5 (27%) | 21.1 (19%) | 28.9 (11%) | 29.9 (20%) | 29.1 (40%) | 45.9 -(35%) | 14.1 (52%) | 21.7 (41%) |
| after 26/38 week home treatm | 17.8 (13%) | 27.6 -(16%) | | 44.2 (16%) | 17.4 (23%) | 14.9 (43%) | 34.4 -(6%) | 34.2 (9%) | 19.8 (59%) | 35.9 -(6%) | 21.1 (28%) | 27.9 (24%) |
| **STEP TEST (number of steps)** | | | | | | | | | | | | |
| **affected foot** | | | | | | | | | | | | |
| 12 weeks prior to treatment | | | | | | | 17 | 11 | 14 | 9 | 9 | 7 |
| immediately before treatment | 12 | 12 | 8 | 14 | 18 | 14 | 21 (24%) | 12 (9%) | 15 (7%) | 13 (44%) | 13 (44%) | 15 (114%) |
| after 4 weeks treatment (CT) | | | | | | | 23 (35%) | 14 (27%) | 17 (21%) | 15 (67%) | 15 (67%) | 14 (100%) |
| after 12 weeks treatment (CT) | 15 (25%) | 18 (50%) | 11 (38%) | 20 (43%) | 23 (28%) | 17 (21%) | 23 (35%) | 14 (27%) | 19 (36%) | 11 (22%) | 16 (78%) | 15 (114%) |
| after 26/38 week home treatm | 15 (25%) | nd | nd | 18 (29%) | 21 (17%) | 17 (21%) | 20 (18%) | 12 (9%) | 21 (50%) | 11 (22%) | 18 (100%) | 14 (100%) |
| **dominant foot** | | | | | | | | | | | | |
| 12 weeks prior to treatment | | | | | | | 12 | 10 | 14 | 11 | 12 | 7 |
| immediately before treatment | 12 | 11 | 9 | 12 | 20 | 14 | 18 (50%) | 12 (20%) | 15 (7%) | 11 (0%) | 12 (0%) | 15 (114%) |
| after 4 weeks treatment (CT) | | | | | | | 21 (75%) | 12 (20%) | 17 (21%) | 15 (36%) | 15 (25%) | 14 (100%) |
| after 12 weeks treatment (CT) | 14 (17%) | 18 (64%) | 11 (22%) | 16 (33%) | 22 (10%) | 17 (21%) | 21 (75%) | 12 (20%) | 19 (36%) | 13 (18%) | 17 (42%) | 15 (114%) |
| after 26/38 week home treatm | 14 (17%) | nd | nd | 20 (67%) | 21 (5%) | 17 (21%) | 24 (100%) | 13 (30%) | 21 (50%) | 13 (18%) | 19 (58%) | 14 (100%) |
| **TS (seconds)** | | | | | | | | | | | | |
| **affected foot behind** | | | | | | | | | | | | |
| 12 weeks prior to treatment | | | | | | | 8.0 | 0.0 | 2.0 | 4.0 | 2.0 | 0.0 |
| immediately before treatment | 30.0 | 12.0 | 0.0 | 0.0 | 1.0 | 6.0 | 8.0 (0%) | 1.0 | 7.5 (275%) | 14.0 (250%) | 3.6 (80%) | 0.0 |
| after 4 weeks treatment (CT) | | | | | | | 30.0 (275%) | 4.7 | 30.0 (1400%) | 30.0 (650%) | 9.0 (350%) | 2.7 |
| after 12 weeks treatment (CT) | 30.0 | 30.0 (150%) | 9.5 | 0.0 | 2.0 (100%) | 2.0 -(67%) | 15.0 (88%) | 1.0 | 5.6 (180%) | 30.0 (650%) | 6.0 (200%) | 1.0 |
| after 26/38 week home treatm | 30.0 | nd | nd | 0.0 | 4.0 (300%) | 3.9 -(34%) | 30.0 (275%) | nd | 30.0 (1400%) | 3.9 -(2%) | 3.4 (70%) | 3.0 |
| **unaffected foot behind** | | | | | | | | | | | | |
| 12 weeks prior to treatment | | | | | | | 1.0 | 0.0 | 1.0 | 7.0 | 4.0 | 0.0 |
| immediately before treatment | 30.0 | 2.0 | 0.0 | 0.0 | 4.0 | 7.0 | 9.2 (820%) | 0.0 | 4.3 (330%) | 4.0 -(43%) | 13.0 (225%) | 0.0 |
| after 4 weeks treatment (CT) | | | | | | | 30.0 (2900%) | 3.1 | 3.0 (200%) | 30.0 (329%) | 3.0 -(25%) | 3.3 |
| after 12 weeks treatment (CT) | 24.3 -(19%) | 30.0 ##### | 2.3 | 1.0 | 3.0 -(25%) | 6.0 -(14%) | 30.0 (2900%) | 2.5 | 2.0 (100%) | 20.0 (186%) | 2.0 -(50%) | 1.0 |
| after 26/38 week home treatm | 30.0 (0%) | nd | nd | 0.0 | 2.7 -(33%) | 3.9 -(44%) | 30.0 (2900%) | nd | 10.7 (972%) | 30.0 (329%) | 3.4 -(15%) | 3.2 |
| **SLS (seconds)** | | | | | | | | | | | | |
| **affected foot raised** | | | | | | | | | | | | |
| 12 weeks prior to treatment | | | | | | | 8.9 | 0.0 | 0.0 | 1.0 | 2.0 | 2.0 |
| immediately before treatment | 6.0 | 2.0 | 5.0 | 0.0 | 8.0 | 1.0 | 2.0 -(78%) | 0.0 | 1.1 | 3.1 (210%) | 2.0 (0%) | 1.0 -(50%) |
| after 4 weeks treatment (CT) | | | | | | | 30.0 (237%) | 0.1 | 1.3 | 1.6 (60%) | 0.0 | 1.0 -(50%) |
| after 12 weeks treatment (CT) | 2.6 -(57%) | 14.0 (600%) | 0.0 | 0.0 | 0.0 | 2.0 (100%) | 10.0 (12%) | 0.0 | 1.6 | 5.4 (440%) | 0.0 | 0.0 |
| after 26/38 week home treatm | 3.9 -(35%) | nd | nd | | 5.4 -(32%) | 1.9 (90%) | 23.2 (161%) | nd | 5.0 | 2.1 (110%) | nd | nd |
| **unaffected foot raised** | | | | | | | | | | | | |
| 12 weeks prior to treatment | | | | | | | 4.9 | 0.0 | 0.0 | 1.0 | 1.0 | 1.0 |
| immediately before treatment | 5.0 | 2.0 | 3.0 | 1.0 | 3.0 | 2.2 | 12.0 (145%) | 0.0 | 2.1 | 3.1 (210%) | 2.0 (100%) | 0.7 -(30%) |
| after 4 weeks treatment (CT) | | | | | | | 6.7 (37%) | 0.1 | 0.1 | 1.6 (60%) | 0.0 | 0.0 |
| after 12 weeks treatment (CT) | 2.4 -(52%) | 2.7 (35%) | 0.0 | 12.3 (1130%) | 0.0 | 1.0 -(55%) | 26.0 (431%) | 0.0 | 1.7 | 5.4 (440%) | 0.0 | 0.0 |
| after 26/38 week home treatm | 18.9 (278%) | nd | nd | | 30.0 (900%) | 30.0 (1264%) | 30.0 (512%) | nd | 5.0 | 2.1 (110%) | nd | nd |
| **WALK SPEED (metres/second)** | | | | | | | | | | | | |
| 12 weeks prior to treatment | | | | | | | 1.22 | 1.03 | 1.62 | 1.20 | 1.33 | 0.76 |
| immediately before treatment | 1.00 | 0.86 | 0.57 | 1.03 | 1.33 | 1.20 | 1.76 (44%) | 1.54 (50%) | 1.88 (16%) | 1.94 (61%) | 1.90 (43%) | 1.50 (98%) |
| after 4 weeks treatment (CT) | | | | | | | 1.94 (58%) | 1.62 (58%) | 2.00 (23%) | 1.94 (61%) | 2.04 (53%) | 1.55 (104%) |
| after 12 weeks treatment (CT) | 1.54 (54%) | 1.62 (89%) | 0.79 (39%) | 1.67 (61%) | 1.94 (45%) | 1.71 (43%) | 1.94 (58%) | 1.60 (56%) | 2.21 (36%) | 1.94 (61%) | 1.96 (47%) | 1.50 (98%) |
| after 26/38 week home treatm | 1.72 (72%) | nd | nd | 1.33 (29%) | 1.75 (32%) | 1.09 -(9%) | 2.07 (69%) | 1.69 (64%) | 2.54 (57%) | 1.93 (61%) | 2.00 (50%) | 1.54 (103%) |
| **WALK STRIDE LENGTH (metres)** | | | | | | | | | | | | |
| 12 weeks prior to treatment | | | | | | | 0.5 | 0.5 | 0.5 | 0.6 | 0.6 | 0.5 |
| immediately before treatment | 0.5 | 0.5 | 0.4 | 0.5 | 0.6 | 0.5 | 0.6 (20%) | 0.7 (33%) | 0.8 (38%) | 0.9 (43%) | 0.6 (0%) | 0.7 (33%) |
| after 4 weeks treatment (CT) | | | | | | | 0.4 -(20%) | 0.8 (50%) | 0.7 (22%) | 0.9 (43%) | 0.8 (25%) | 0.7 (33%) |
| after 12 weeks treatment (CT) | 0.7 (22%) | 0.5 (18%) | 0.5 (8%) | 0.7 (33%) | 0.5 -(23%) | 0.8 (38%) | 0.7 (33%) | 0.7 (33%) | 0.8 (38%) | 0.9 (43%) | 0.8 (25%) | 0.7 (33%) |
| after 26/38 week home treatm | 0.8 (38%) | nd | nd | 0.6 (20%) | 0.8 (25%) | 0.7 (22%) | 0.8 (50%) | 0.7 (33%) | 0.8 (38%) | 0.8 (25%) | 0.8 (25%) | 0.8 (50%) |
| **TUG (seconds)** | | | | | | | | | | | | |
| 12 weeks prior to treatment | | | | | | | 7.9 | 9.7 | 6.8 | 7.6 | 7.8 | 13.8 |
| immediately before treatment | 10.2 | 8.0 | 18.0 | 11.0 | 6.5 | 7.5 | 7.4 (6%) | 8.5 (12%) | 6.5 (4%) | 7.3 (4%) | 6.7 (14%) | 9.3 (33%) |
| after 4 weeks treatment (CT) | | | | | | | 6.4 (19%) | 7.4 (24%) | 6.3 (7%) | 6.6 (13%) | 6.5 (17%) | 7.9 (43%) |
| after 12 weeks treatment (CT) | 8.4 (18%) | 7.1 (11%) | 15.5 (14%) | 8.5 (23%) | 5.2 (20%) | 6.6 (12%) | 6.6 (16%) | 7.6 (22%) | 5.9 (13%) | 7.1 (6%) | 6.3 (19%) | 7.6 (45%) |
| after 26/38 week home treatm | 8.1 (21%) | nd | nd | 9.9 (10%) | 6.0 (7%) | 5.8 (23%) | 6.6 (16%) | 8.0 (18%) | 6.3 (7%) | 6.6 (13%) | 5.9 (24%) | 7.9 (43%) |
| **TUG MOTOR (seconds)** | | | | | | | | | | | | |
| 12 weeks prior to treatment | | | | | | | 8.8 | 9.0 | 6.9 | 7.6 | 8.3 | 11.8 |
| immediately before treatment | 10.9 | 8.2 | 17.7 | 10.6 | 7.1 | 7.2 | 8.2 (7%) | 8.1 (10%) | 6.5 (6%) | 8.3 -(9%) | 6.4 (23%) | 9.1 (23%) |
| after 4 weeks treatment (CT) | | | | | | | 7.4 (16%) | 7.2 (20%) | 6.6 (4%) | 6.9 (9%) | 6.4 (23%) | 8.3 (30%) |
| after 12 weeks treatment (CT) | 8.8 (19%) | 8.2 (0%) | 16.5 (7%) | 9.3 (12%) | 6.6 (7%) | 7.0 (3%) | 6.4 (27%) | 8.5 (6%) | 6.3 (9%) | 7.1 (7%) | 6.6 (20%) | 8.1 (31%) |
| after 26/38 week home treatm | 8.4 (23%) | nd | nd | 13.3 -(26%) | 6.1 (14%) | 6.4 (11%) | 6.0 (32%) | 9.2 -(2%) | 7.0 -(1%) | 6.8 (10%) | 5.8 (30%) | 8.4 (29%) |
| **TUG COGNITIVE (seconds)** | | | | | | | | | | | | |
| 12 weeks prior to treatment | | | | | | | 10.3 | 10.6 | 7.2 | 8.7 | 8.8 | 13.8 |
| immediately before treatment | 12.0 | 10.4 | 23.7 | 10.5 | 6.0 | 7.6 | 7.5 (27%) | 8.8 (17%) | 7.3 -(1%) | 7.5 (14%) | 6.7 (24%) | 9.0 (35%) |
| after 4 weeks treatment (CT) | | | | | | | 7.2 (30%) | 8.2 (23%) | 6.7 (7%) | 6.6 (24%) | 6.2 (30%) | 8.7 (37%) |
| after 12 weeks treatment (CT) | 9.3 (23%) | 6.9 (34%) | 18.2 (23%) | 9.3 (11%) | 5.2 (14%) | 6.8 (11%) | 6.3 (39%) | 8.9 (16%) | 6.4 (11%) | 6.8 (22%) | 6.8 (23%) | 8.1 (41%) |
| after 26/38 week home treatm | 8.8 (26%) | nd | nd | 9.8 (6%) | 5.2 (13%) | 5.9 (22%) | 6.7 (35%) | 9.5 (11%) | 6.5 (10%) | 6.4 (27%) | 6.6 (25%) | 8.8 (36%) |
| MoCA score | | | | | | | | | | | | |
| 12 weeks prior to treatment | | | | | | | 26 | 28 | 27 | 26 | 24 | 21 |
| immediately before treatment | 24 | 26 | 26 | 28 | 27 | 24 | 29 (12%) | 29 (4%) | 27 (0%) | 26 (0%) | 28 (17%) | 24 (14%) |
| after 4 weeks treatment (CT) | | | | | | | 28 (8%) | 30 (7%) | 28 (4%) | 26 (0%) | 28 (17%) | 25 (19%) |
| after 12 weeks treatment (CT) | 25 (4%) | 26 (0%) | 28 (8%) | 28 (0%) | 27 (0%) | 29 (21%) | 29 (12%) | 30 (7%) | 27 (0%) | 27 (4%) | 29 (21%) | 29 (38%) |
| after 26/38 week home treatm | 29 (21%) | 29 (12%) | nd | 30 (7%) | 28.7 (6%) | 30 (24%) | 30 (15%) | 30 (7%) | 30 (11%) | 28.7 (10%) | 30 (25%) | nd |

**NHPT = nine-hole peg test**
**TS = tandem stance (eyes closed)**
**SLS = single leg stance (eyes closed)**
**TUG = timed up-and-go**
**MoCA = Montreal Cognitive Assessment**
**number in brackets is the percentage improvement (positive value) or decline (negative value) in an outcome measure**
